# Epidemiology of acute respiratory infections in adult patients in Hospices Civils de Lyon, France, between October 2016 and April 2025

**DOI:** 10.64898/2026.11.04.26367246

**Authors:** Cécile Chauvel, Cédric Dananché, George Shirreff, Claire Nour Abou Chakra, Alexandre Gaymard, Philippe Vanhems, Florence Morfin, Laurence Josset, Antonin Bal, Jean-Sebastien Casalegno, Marta C Nunes

## Abstract

**Background:** Multiple viruses can cause acute respiratory infections (ARI), impacting morbidity, mortality and hospital resources. The objective of this study was to describe viral circulation among patients with ARI, patients’ characteristics and severity outcomes.

**Methods:** A retrospective cohort was assembled using electronic health records from the Hospices Civils de Lyon, France. Adult patients (≥18 years) hospitalised or presenting to the emergency department from October 2016 to April 2025 with either an ICD-10 diagnostic code for ARI or a laboratory-confirmed viral detection were included. Administrative variables and laboratory results collected during the stay were extracted.

**Results:** 103,011 hospital visits involving 82,077 patients were included. The number of eligible stays increased substantially since the COVID-19 pandemic, along with year-round laboratory testing. Following disruption during the pandemic, winter peaks of ARI incidence, influenza and RSV detections re-emerged after 2022. Influenza, rhinovirus and SARS-CoV-2 positivity rates peaked in younger adults (<65 years), whereas RSV and hMPV positivity rates were highest in patients aged ≥80 years. Patients aged ≥65 years had longer hospitalisations (median 10 vs 8 days, p<0.001), twice in-hospital mortality rate (14% vs 7%, p<0.001), less frequent ICU admission (21% vs 33%, p<0.001) and use of respiratory support than younger adults. Influenza was the virus associated with the shortest length of stays, hMPV was associated with the longest in younger adults.

**Conclusions:** Real-world hospital data are valuable for surveillance of viral circulation, early identification of patients at risk of deterioration, and may be used to evaluate the impact of preventive intervention in community.

## INTRODUCTION

Acute respiratory infections (ARIs) remain a major cause of morbidity, mortality and healthcare utilisation worldwide, particularly during seasonal epidemics and emerging respiratory virus outbreaks^1^. Hospital-based surveillance plays a central role in monitoring the circulation of respiratory pathogens, evaluating disease severity and informing public health interventions^2,3^. The COVID-19 pandemic further highlighted the importance of timely surveillance systems capable of tracking changes in respiratory virus epidemiology, healthcare demand and diagnostic practices^4^.

Electronic health records provide an increasingly important source of real-world data for respiratory surveillance because they capture large numbers of patients managed under routine clinical conditions. However, existing studies have generally relied either on administrative databases, which identify cases using diagnostic codes, or on prospective surveillance cohorts with systematic virological testing in selected populations or predefined surveillance periods^5–8^. While each approach has important strengths, few studies have integrated administrative, clinical and laboratory data within the same healthcare system over an extended period, limiting the ability to jointly describe healthcare utilisation, respiratory virus circulation, patient characteristics and severity outcomes in routine practice^9^.

The objective of this study was to describe respiratory virus circulation including temporal trends in hospital activity related to ARI, patients’ characteristics and severity outcomes in the longitudinal cohort of adult patients with ARI attending the Hospices Civils de Lyon (HCL) from October 2016 to April 2025.

## METHODS

### Study design and data source

We conducted a retrospective cohort study using routinely collected electronic health record data from HCL, the second largest university hospital in France, serving the city of Lyon and its surrounding metropolitan area (approximately 1.4 million inhabitants)^10^. Administrative records, severity outcomes and laboratory test results generated during routine patient care were linked at the individual hospital-visit level and pseudonymised prior to analysis. Eligible patients were adults (≥18 years) who visited an HCL emergency department (ED) or were hospitalised with an ARI from October 1^st^, 2016 to April 30^th^, 2025. Patients were included if they had either (i) a laboratory-confirmed detection of influenza virus, respiratory syncytial virus (RSV), severe acute respiratory syndrome coronavirus 2 (SARS-CoV-2), human metapneumovirus (hMPV), rhinovirus/enterovirus (RV/EV), adenovirus (ADV), parainfluenza virus (PIV), or human coronavirus (hCoV), or (ii) at least a diagnostic code corresponding to ARI according to the *International Classification of Diseases, Tenth Revision* (ICD-10) at discharge either from hospital or from any ward during their stay (Supplementary Table 1). The study covered nine respiratory virus seasons, with epidemic phases considered from October 1^st^ to April 30^th^ and interseason periods from May 1^st^ to September 30^th^ of each year. Data for the final 2024-2025 period were available until April 30^th^, 2025. Virological tests did not always enable to distinguish between rhinovirus and enterovirus. From November 2018, Panther Fusion® respiratory virus assays specific to rhinovirus was implemented at HCL^11^. Tests for RV/EV or specific to rhinovirus were used subsequently, based on the clinicians’ decision.

### Variables and outcome definitions

Administrative variables comprised age, sex, department of residence, date of admission, discharge status (in-hospital death, home or transfer to another healthcare facility), type of hospital visit (hospitalisation or ED visit not followed by a hospitalisation) and ICD-10 diagnostic codes. Comorbidities were identified from ICD-10 codes assigned during the ARI hospitalisation (Supplementary Table 1). Severity outcomes included length of hospitalisation in days, in-hospital death, admission to an intensive care unit (ICU), use of oxygen supplementation, non-invasive mechanical ventilation (non-IMV), invasive mechanical ventilation (IMV) or extra-corporeal membrane oxygenation (ECMO). Laboratory variables included results of virological tests performed during the stay, for any of the viruses used for patient inclusion. For each hospital visit, multiple virological test results for the same virus were collapsed into a single result, classified as positive if at least one test was positive and negative if all were negative. All influenza-positive samples were typed; untyped samples were excluded (n=46, all collected before the 2019-2020 season). Consecutive hospital readmissions occurring within 30 days were considered related to the same hospital visit. Comorbidities, ICU admission and use of respiratory support were documented from any of these consecutive readmissions within 30 days. The discharge outcome (home, transfer in another health care setting or in-hospital death) of the final visit was retained, whereas the duration of the first admission was used when analysing length of hospitalisation.

### Data analysis

The number of ED visits and hospitalisations included through laboratory-confirmed viral detection or ICD-10 coding were summarised by respiratory virus season. Visits meeting both inclusion criteria were classified according to laboratory confirmation, which occurred before the diagnostic coding.

Monthly incidence rates of ARI-related hospital visits per 100,000 population were calculated by age group and type of hospital visit using previously estimated HCL catchment populations^12^. Catchment populations were derived using a proportional patient-flow approach based on population data from *Institut National de la Statistique et des Etudes Economiques (INSEE)*, hospitalisation data from *Agence Technique de l’Information sur l’Hospitalisation (ATIH)*, and, since 2019, ED attendance data from the *Réseau d’Urgences d’Auvergne-Rhône-Alpes* (Urg’ARA), stratified by age, calendar year and postcode of residence. Since Urg’ARA started the data collection in 2019, the numbers of patients from the HCL catchment area visiting the ED in 2019 were used for the preceding seasons. Monthly 95% confidence intervals (95% CI) were also reported^13^.

Temporal patterns of respiratory virus circulation were described for each virus by season and month using the number of documented laboratory-confirmed detections. Positivity rates in patients with ARI were calculated by age group and season for each virus. The distribution of tested respiratory virus across age groups was also described for each season.

Patient characteristics were summarised separately for hospitalisations and ED-only visits according to sex, age, residence in the Rhône department (the same department as the HCL), readmissions within 30 days and period relative to the COVID-19 pandemic (pre-pandemic: 2016-2017 to 2018-2019; pandemic: 2019-2020 to 2021-2022; post-pandemic: 2022-2023 to 2024-2025). For hospitalised patients, ED attendance during the stay, number of comorbidities (none, one, or at least two), and individual comorbidities were also described. Cardiovascular diseases were reported both including and excluding hypertension because of its high prevalence in older adults^14–16^. Characteristics, laboratory-confirmed viral detection and severity outcomes were additionally summarized by age group (18–64, 65–79 and ≥80 years). For patients hospitalised after October 1^st^, 2020, length of stay and oxygen supplementation were compared between those with laboratory-confirmed influenza A, influenza B, SARS-CoV-2, RSV, hMPV or rhinovirus detection. Patients with laboratory-confirmed viral co-detections were excluded. Analyses were stratified by age group (18-64 and ≥65 years) and by ICU admission status, and ventilation interventions were compared only for stays including an admission to ICU. Statistical testing involving influenza B in patients aged ≥65 years were not performed because of the small number of cases.

Chi-square tests were used to compare categorical variables and either Wilcoxon (for 2-category outcomes) or Kruskal-Wallis (for 3-category outcomes) tests were used for continuous variables. P-values were adjusted for multiple comparisons using the Benjamini-Hochberg procedure, with adjusted p-values <0.05 considered statistically significant^17^. Description of severity outcomes for the three age groups are provided in the supplementary material.

All statistical analyses were conducted using R version 4.4.3^18^.

## RESULTS

### Cohort description

Overall, 103,011 hospital visits involving 82,077 distinct patients with ARI were identified from October 2016 to April 2025, including 19,948 ED visits without hospitalisation and 83,063 hospitalisations (Figure 1 and Supplementary Table 2). Across the study period, most patients were identified through ICD-10 diagnostic codes rather than laboratory-confirmed viral detection, with 59% of ED visits and 64% of hospitalisations lacking in-hospital virological confirmation.

**Figure 1:**
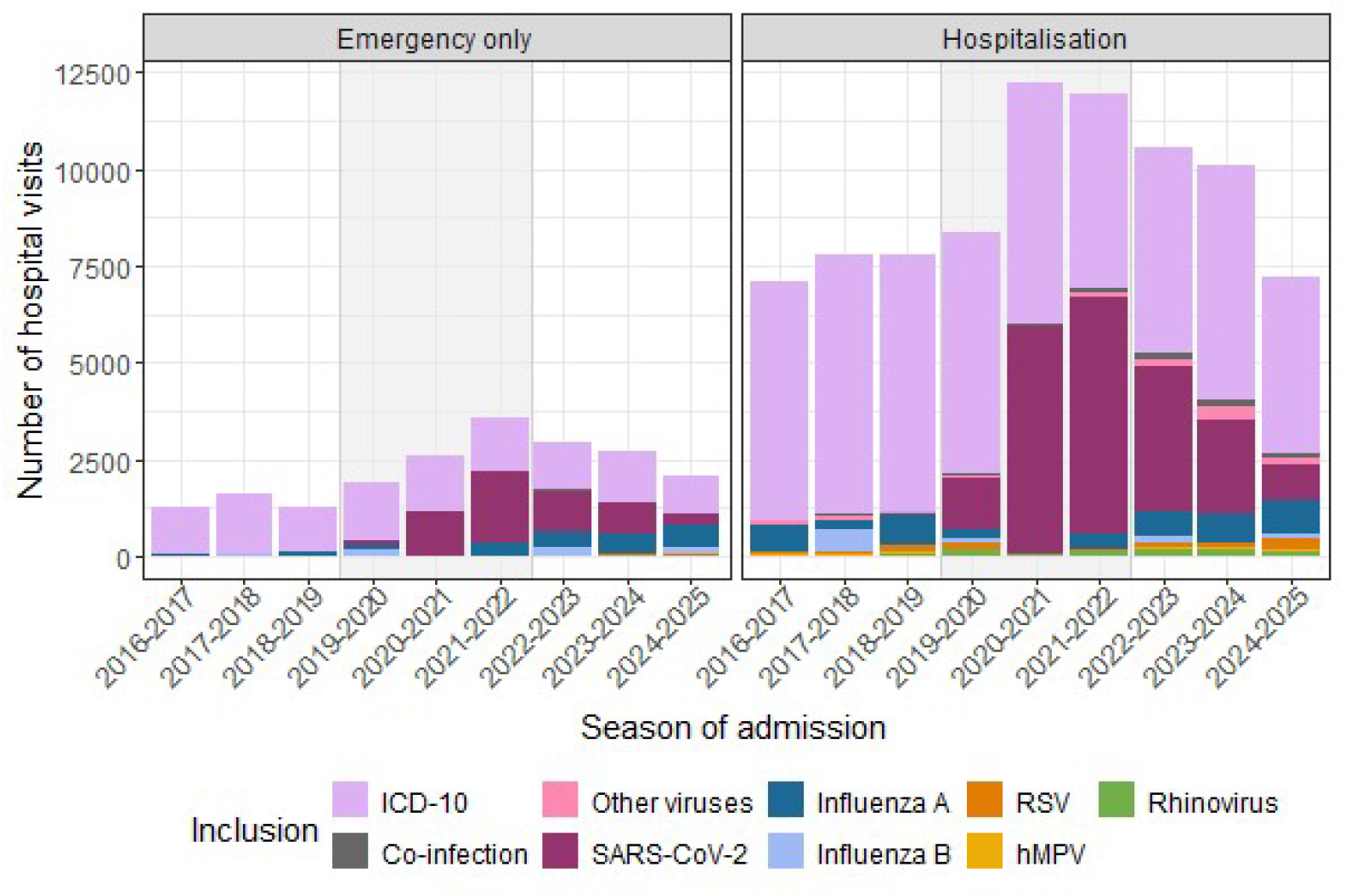
Number of hospital visits by ICD-10 code or laboratory-confirmed viral infection. Grey shading: COVID-19 pandemic seasons. Note: 2024-2025 only covers September 2024 through April 2025

The number of ARI-related hospital visits increased markedly during the COVID-19 pandemic. Compared with the 2018–2019 season, hospitalisations increased by 57% in 2020-2021 and remained 53% higher in 2021-2022, while ED visits increased 1.5-fold in 2019-2020, twofold in 2020–2021 and almost threefold in 2021–2022. After October 2022, the number of ARI hospital visits remained above pre-COVID-19 pandemic levels, with approximately 30% more hospitalisations and twice as many ED visits than before the pandemic.

The proportion of ARI visits with a laboratory-confirmed viral detection increased over time. Among ED visits, laboratory-confirmed cases increased from 8.7% in 2018–2019 to 62% in 2021–2022, while for hospitalisations they increased from 15% to 58% over the same period. After October 2021, ED visits surpassed hospitalisations in terms of the proportion of visits with laboratory confirmation.

### Incidence of ARI hospital visits

Monthly incidence rates in the HCL catchment population showed marked seasonal peaks before the COVID-19 pandemic, particularly for hospitalisations in older adults (Figure 2). During the pandemic, incidence patterns changed substantially across all age groups. Hospitalisation incidences in adults aged 18-64 years and 65-79 years increased from 2020-2021 before declining after October 2022, although remaining above pre-pandemic levels. Historical seasonal patterns were observed again after the COVID-19 pandemic. In contrast, ED visits incidence in adults aged ≥65 years increased progressively throughout the pandemic and remained persistently high thereafter.

**Figure 2:**
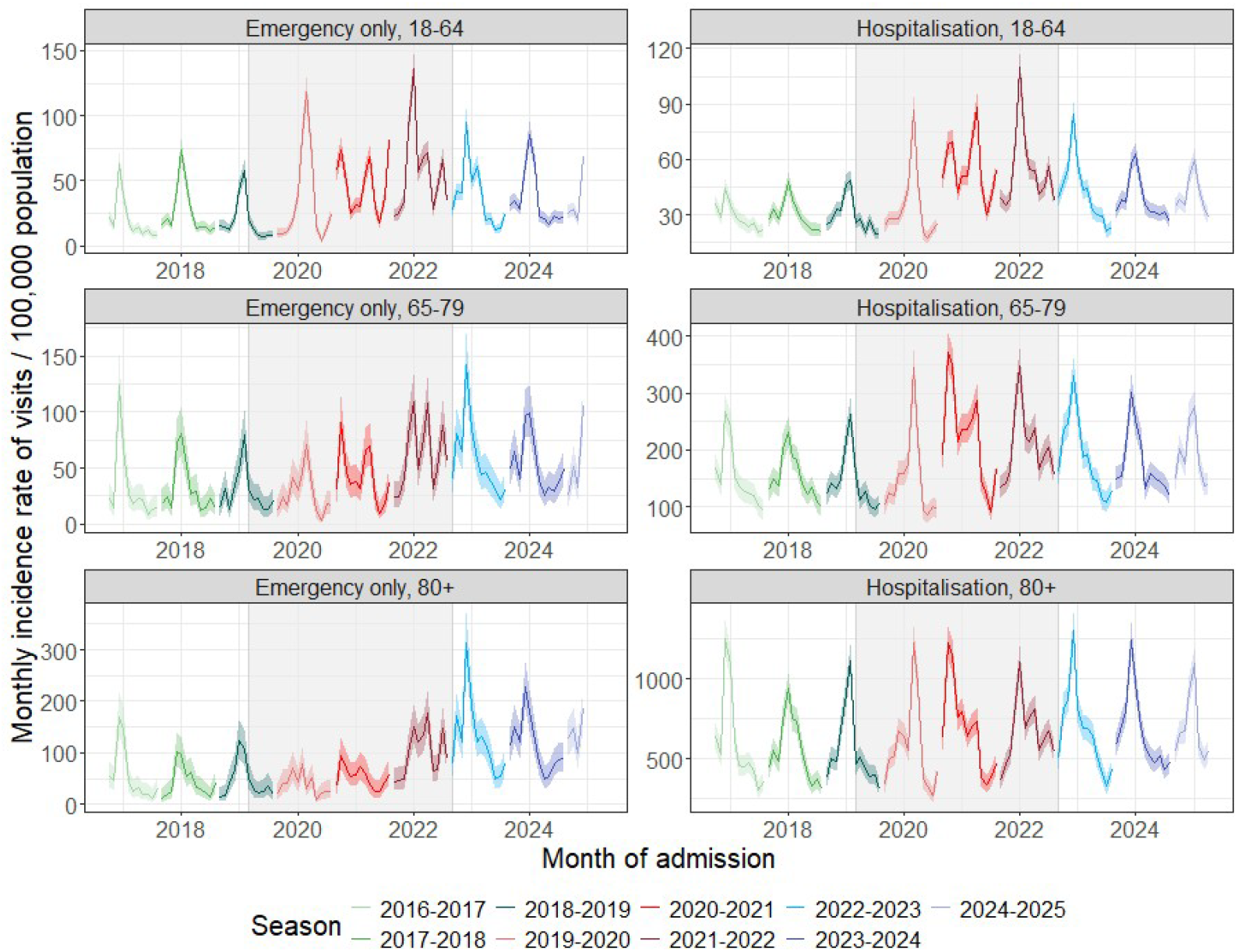
Monthly incidence rates (curves) and 95% confidence intervals (bands) of hospital visits with acute respiratory infection by type of visit and age group in the catchment area of the Hospices Civils de Lyon. Grey shading: COVID-19 pandemic seasons. Y-axis scales differ by panel

### Virological testing and virus circulation

The COVID-19 pandemic was associated with major changes in respiratory virus testing and detection (Figure 3; Supplementary Figures 1 and 3). During the 2020-2021 season, influenza and most other respiratory viruses were rarely detected, whereas SARS-CoV-2 accounted for the majority of laboratory-confirmed detections. From 2022 onwards, both the number of respiratory virus tests and the number of laboratory-confirmed detections increased substantially for all viruses except influenza B, which continued to display high circulation every other year.

**Figure 3:**
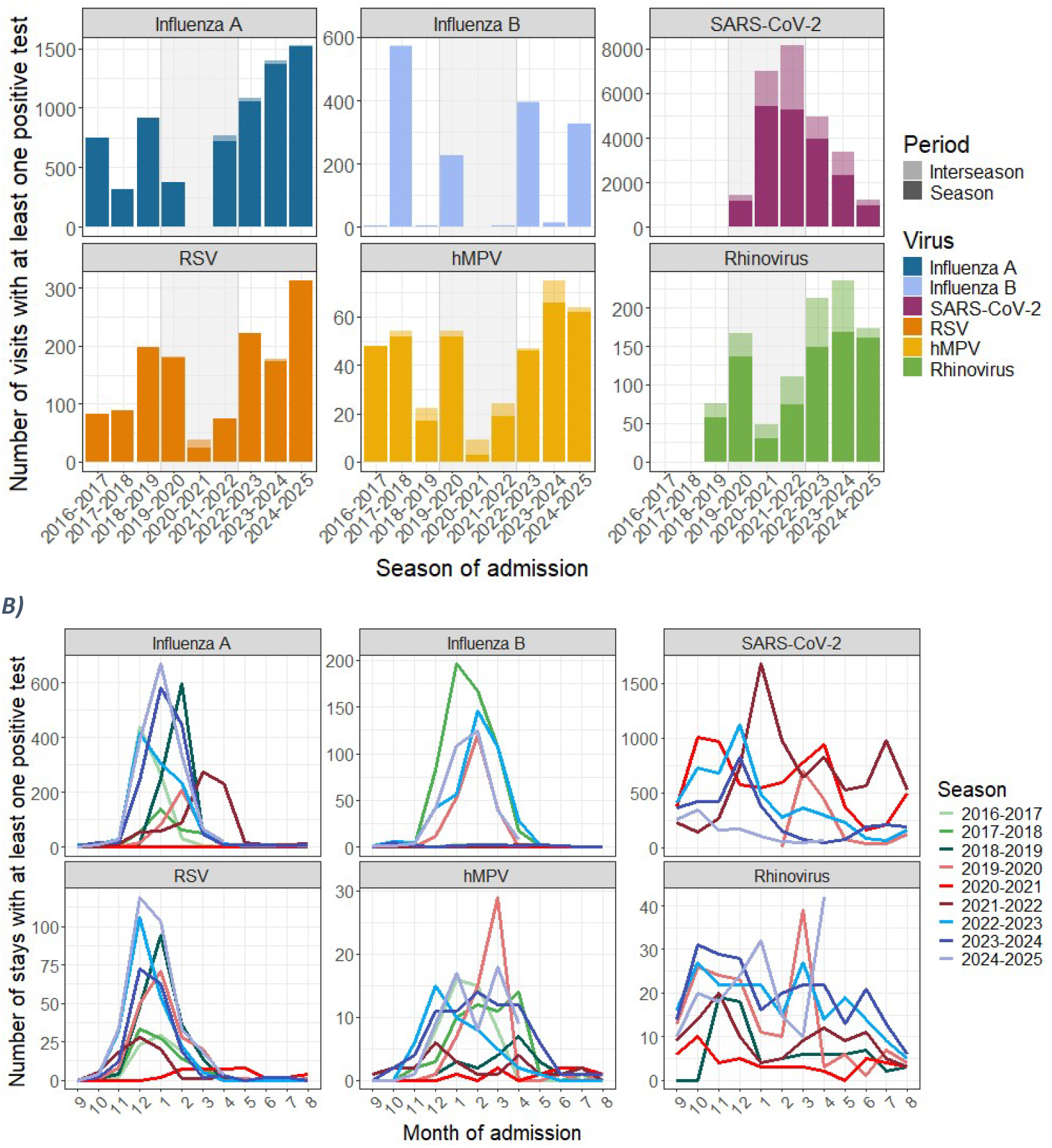
Number of hospital visits with at least one positive viral test for influenza A, influenza B, SARS-CoV-2, RSV, hMPV and rhinovirus by epidemic season (Figure 3A) and month (Figure 3B) Grey shading: COVID-19 pandemic seasons. 2024-2025 only covers September 2024 through April 2025. Y-axis scales differ by panel

Testing outside the traditional respiratory virus season became common after the COVID-19 pandemic for all viruses (Supplementary Figure 1). Despite this expansion in testing, influenza and RSV continued to exhibit well-defined winter epidemic seasonality with very few detections between May and September (Figure 3B). hMPV cases were generally detected from November to May, whereas SARS-CoV-2 and rhinovirus displayed less distinct seasonal patterns.

Positivity rates differed by age group (Supplementary Figure 2). Since the 2019-2020 season, influenza, rhinovirus and SARS-CoV-2 positivity rates were generally highest in adults aged 18–64 years, whereas RSV and hMPV positivity rates were highest in adults aged ≥80 years, except during the 2020–2021 season when low circulation of these viruses.

### Characteristics of patients with ARI

Most hospital visits involved patients residing in the Rhône department, at a stable rate over time (Table 1). Patients visiting the ED without hospitalisation were younger than those hospitalised (median age 49 vs 73 years, p<0.001), and women accounted for a greater proportion of ED visits than hospitalisations (54% vs 45%, p<0.001). An ED visit was observed in 60% of hospitalisations, with this proportion increasing with the pandemic (55% before vs 62% after, p<0.001). The proportion of patients with one comorbidity remained relatively stable over time (23-24%, p<0.001), while the proportion with multiple comorbidities declined during the pandemic (from 59% to 54%, p<0.001). The most frequent comorbidities were cardiovascular diseases (47%), even when excluding hypertension (31%), chronic lung disease (21%) and diabetes (20%) (Supplementary Table 3).

**Table 1:**
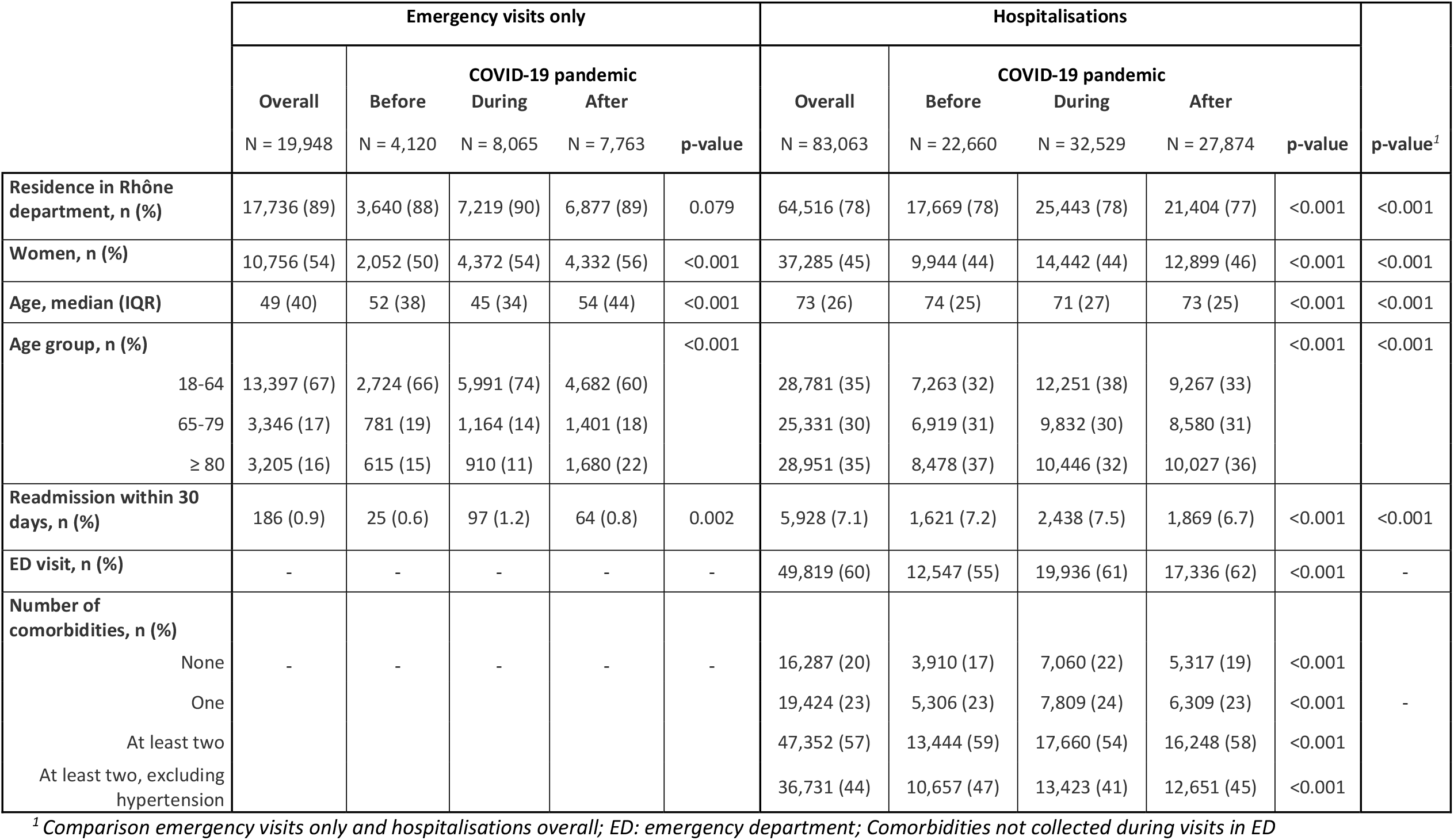
Characteristics of patients with acute respiratory infection by type of hospital visit, overall and by period related to the COVID-19 pandemic.

|  | **Emergency visits only** |  |  |  |  | **Hospitalisations** |  |  |  |  |  |
|---|---|---|---|---|---|---|---|---|---|---|---|
|  |  | **COVID-19 pandemic** |  |  |  |  | **COVID-19 pandemic** |  |  |  |  |
|  | **Overall** | **Before** | **During** | **After** |  | **Overall** | **Before** | **During** | **After** |  |  |
|  | N = 19,948 | N = 4,120 | N = 8,065 | N = 7,763 | **p-value** | N = 83,063 | N = 22,660 | N = 32,529 | N = 27,874 | **p-value** | **p-value<sup>*1*</sup>** |
| **Residence in Rhône department, n (%)** | 17,736 (89) | 3,640 (88) | 7,219 (90) | 6,877 (89) | 0.079 | 64,516 (78) | 17,669 (78) | 25,443 (78) | 21,404 (77) | <0.001 | <0.001 |
| **Women, n (%)** | 10,756 (54) | 2,052 (50) | 4,372 (54) | 4,332 (56) | <0.001 | 37,285 (45) | 9,944 (44) | 14,442 (44) | 12,899 (46) | <0.001 | <0.001 |
| **Age, median (IQR)** | 49 (40) | 52 (38) | 45 (34) | 54 (44) | <0.001 | 73 (26) | 74 (25) | 71 (27) | 73 (25) | <0.001 | <0.001 |
| **Age group, n (%)** |  |  |  |  | <0.001 |  |  |  |  | <0.001 | <0.001 |
| 18-64 | 13,397 (67) | 2,724 (66) | 5,991 (74) | 4,682 (60) |  | 28,781 (35) | 7,263 (32) | 12,251 (38) | 9,267 (33) |  |  |
| 65-79 | 3,346 (17) | 781 (19) | 1,164 (14) | 1,401 (18) |  | 25,331 (30) | 6,919 (31) | 9,832 (30) | 8,580 (31) |  |  |
| ≥ 80 | 3,205 (16) | 615 (15) | 910 (11) | 1,680 (22) |  | 28,951 (35) | 8,478 (37) | 10,446 (32) | 10,027 (36) |  |  |
| **Readmission within 30 days, n (%)** | 186 (0.9) | 25 (0.6) | 97 (1.2) | 64 (0.8) | 0.002 | 5,928 (7.1) | 1,621 (7.2) | 2,438 (7.5) | 1,869 (6.7) | <0.001 | <0.001 |
| **ED visit, n (%)** | - | - | - | - | - | 49,819 (60) | 12,547 (55) | 19,936 (61) | 17,336 (62) | <0.001 | - |
| **Number of comorbidities, n (%)** |  |  |  |  |  |  |  |  |  |  |  |
| None | - | - | - | - | - | 16,287 (20) | 3,910 (17) | 7,060 (22) | 5,317 (19) | <0.001 |  |
| One |  |  |  |  |  | 19,424 (23) | 5,306 (23) | 7,809 (24) | 6,309 (23) | <0.001 | - |
| At least two |  |  |  |  |  | 47,352 (57) | 13,444 (59) | 17,660 (54) | 16,248 (58) | <0.001 |  |
| At least two, excluding hypertension |  |  |  |  |  | 36,731 (44) | 10,657 (47) | 13,423 (41) | 12,651 (45) | <0.001 |  |

<sup>*1*</sup> *Comparison emergency visits only and hospitalisations overall; ED: emergency department; Comorbidities not collected during visits in ED*

**Table 2:** Characteristics of patients hospitalised with acute respiratory infection and their severity outcomes overall and by age group.

|  |  | Age group |  |  |
|---|---|---|---|---|
|  | **Overall**<br>N = 83,063<sup>*1*</sup> | **18-64**<br>N = 28,781<sup>*1*</sup> | **≥65**<br>N = 54,2821<sup>*1*</sup> | **p-value** |
| **Length of first stay in days, median (IQR)** | 9 (20) | 8 (25) | 10 (17) | <0.001 |
| **Readmission within 30 days, n (%)** | 5,928 (7.1) | 2,004 (7.0) | 3,924 (7.2) | 0.2 |
| **Women, n (%)** | 37,285 (45) | 12,318 (43) | 24,967 (46) | <0.001 |
| **Emergency department visit, n (%)** | 49,819 (60) | 13,501 (47) | 36,318 (67) | <0.001 |
| **Type of discharge, n (%)** |  |  |  | <0.001 |
| In-hospital death | 9,708 (12) | 1,973 (6.9) | 7,735 (14) |  |
| Home | 51,117 (62) | 20,940 (73) | 30,177 (56) |  |
| Transfer | 22,238 (27) | 5,868 (20) | 16,370 (30) |  |
| **Laboratory-confirmed viral infection, n (%)** | 30,135 (36) | 10,955 (38) | 19,180 (35) | <0.001 |
| **Laboratory-confirmed viral co-infection, n (%)** | 781 (0.9) | 361 (1.3) | 420 (0.8) | <0.001 |
| **Number of comorbidities, n (%)** |  |  |  | <0.001 |
| None | 16,287 (20) | 8,574 (30) | 7,713 (14) |  |
| One | 19,424 (23) | 8,741 (30) | 10,683 (20) |  |
| At least two | 47,352 (57) | 11,466 (40) | 35,886 (66) |  |
| At least two, excluding hypertension | 36,731 (44) | 9,613 (33) | 27,118 (50) |  |
| **ICU, n (%)** | 21,094 (25) | 9,617 (33) | 11,477 (21) | <0.001 |
| **Oxygen supplementation, n (%)** | 16,617 (20) | 7,458 (26) | 9,159 (17) | <0.001 |
| **Non-invasive mechanical ventilation, n (%)** | 12,157 (15) | 5,164 (18) | 6,993 (13) | <0.001 |
| **Invasive mechanical ventilation, n (%)** | 10,283 (12) | 5,269 (18) | 5,014 (9.2) | <0.001 |
| **ECMO, n (%)** | 310 (0.4) | 261 (0.9) | 49 (<0.1) | <0.001 |

*ICU: intensive care unit; ECMO: extracorporeal membrane oxygenation*

### Severity outcomes

Hospitalised patients aged ≥65 years had longer hospital stays than younger adults (median 10 vs 8 days, p<0.001), although 30-day readmission rates were similar across age groups (approximately 7%, p=0.2) (Table2) Older adults were more likely to attend the ED (67% vs 47%, p<0.001), had higher frequencies of multiple comorbidities (66% vs 40%, p<0.001), in-hospital mortality rate twice as high, and were transferred to other healthcare facilities 1.5-times more often (p<0.001). In contrast, younger adults (aged 18-64 years) were more frequently admitted to ICU (33% vs 21%, p<0.001) and more commonly received oxygen supplementation (25% vs 16%, p<0.001), non-IMV (18% vs 13%, p<0.001), IMV (18% vs 9%, p<0.001) and ECMO (0.9% vs <0.1%, p<0.001). Age stratification of older adults showed that patients aged 65–79 years generally had severity outcomes more similar to younger adults than to those aged ≥80 years (Supplementary Table 4).

### Severity according to respiratory virus

For both age groups with confirmed viral detection, hospitalisations after October 2020 lasted longer when ICU stay was required (Figure 4). Influenza detection was generally associated with shorter hospitalisation (for Influenza A, median duration of 4 days for patients aged 18-64 years and 8 days for those aged ≥65 years without ICU) than SARS-CoV-2 (median 5 and 10 days), RSV and rhinovirus (median 6 and 10 days) and hMPV (median 7 and 10.5). In younger adults, detection of hMPV, followed by rhinovirus, and RSV in absence of ICU admission, were generally associated with longer stays.

**Figure 4:**
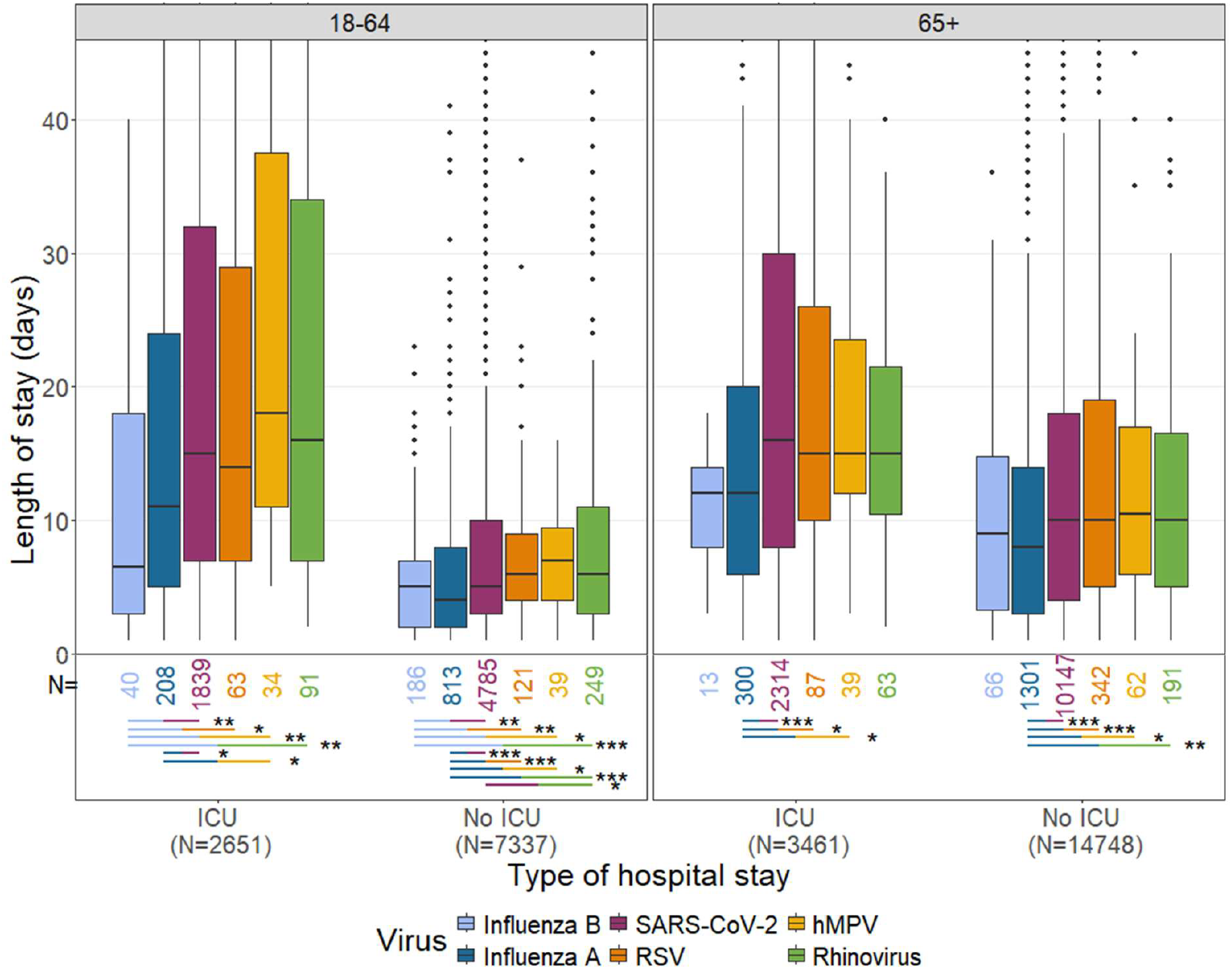
Length of stay for hospitalisations from October 2020 with a positive viral test for influenza A, influenza B, SARS-CoV-2, RSV, hMPV and rhinovirus, by admission to intensive care unit (ICU) and age group. The boxplots present the median (horizontal bar) and inter-quartile range (boxes) of length of hospital stay truncated at 45 days. N represents the number of stays for each viral detection, age group and ICU/non-ICU admission. The horizontal bars below the boxplots display pairwise statistical significance, with the following meaning ***: adj-p <0.001; **: 0.001≤ adj-p <0.01; *: 0.01≤ adj-p <0.05.

For patients admitted to ICU after October 2020, the use of respiratory support depended on the nature of the detected virus (Figure 5). Oxygen supplementation and non-IMV were more frequent in inpatients with RSV or hMPV detections in both age groups. In young adults, rhinovirus detection was consistently associated with lower use of oxygen supplementation (33% of stays) and ventilatory support (27% for non-IMV and 18% for IMV) than influenza A, SARS-CoV-2, RSV or hMPV detection (52-65% for oxygen support, 42-60% for non-IMV and 34-56% for IMV). In adults aged ≥65 years, RSV detection was associated with higher use of oxygen supplementation (81%) and non-IMV (74%) than detection of influenza A, SARS-CoV-2 and rhinovirus (52-60% for oxygen supplementation, 35-58% for non-IMV), whereas IMV was most frequently required in patients with either RSV, hMPV or SARS-CoV-2 detection (37-40%).

**Figure 5:**
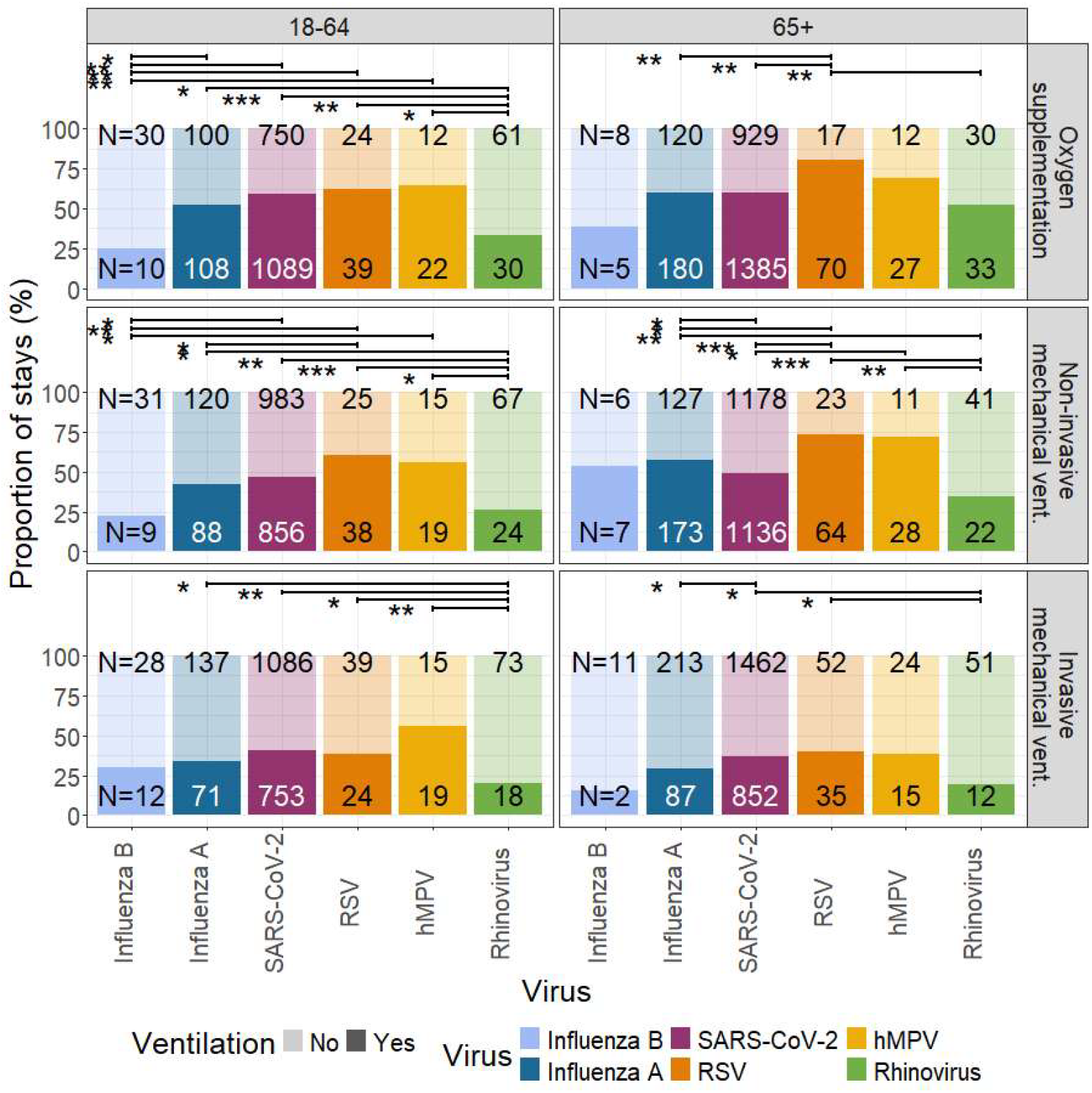
Proportion of use of oxygen supplementation, non-invasive and invasive mechanical ventilations for hospitalisations including an admission to intensive care unit after October 2020, with a positive viral test for influenza A, influenza B, SARS-CoV-2, RSV, hMPV and rhinovirus, by ventilation interventions and age group. N represents the number of stays with ventilation support (bottom) or not (top) for each viral detection and age group. The horizontal bars above the boxplots display pairwise statistical significance, with the following meaning ***: adj-p <0.001; **: 0.001≤ adj-p <0.01; *: 0.01≤ adj-p <0.05

## DISCUSSION

This study describes a large longitudinal cohort of adults presenting with ARI to a major tertiary university hospital over nine consecutive respiratory virus seasons. While large real-world studies have successfully used administrative healthcare data to investigate respiratory infections, they have generally relied on diagnostic coding to identify pathogen-specific hospitalisation^6,7^. Conversely, prospective studies incorporating systematic virological testing have usually been limited to specific viruses, selected populations or predefined surveillance periods, often restricted to the winter^19–22,8^. By linking routinely collected administrative data with laboratory-confirmed detections, the cohort in this study captures the main respiratory viruses within the second largest hospital in France.

The COVID-19 pandemic fundamentally changed both respiratory virus epidemiology and hospital diagnostic practices. In addition to the emergence of SARS-CoV-2, the widespread implementation of molecular diagnostics and the expansion of respiratory virus testing beyond the traditional winter season has led to a large increase in the number of laboratory-confirmed respiratory detections^23^. Consequently, temporal increases in virus detections after 2020 should not be interpreted only as evidence of changing virus circulation. These findings illustrate an important consideration when interpreting routinely collected hospital data: observed temporal changes may result from evolving testing strategies as well as real epidemiological trends. Similar observations have been reported for RSV, where linkage of microbiological and administrative data substantially improved case ascertainment^24^, while adjustment for under-ascertainment has been estimated to more than double RSV hospitalisation rates in older adults^25^.

Despite these changes in testing practices, the epidemiological patterns observed in our study were consistent with the established behaviour of respiratory viruses in temperate climates. Influenza and RSV maintained pronounced winter seasonality, whereas SARS-CoV-2 and rhinovirus showed less clearly defined seasonal patterns^26,27^. hMPV circulated predominantly between late autumn and spring, and together with RSV showed the highest positivity rates in adults ≥80 years, consistent with previous reports describing the burden of these viruses in older adults^20,28–30^. Influenza and rhinovirus positivity rates, in contrast, were generally highest in younger adults. Excluding the pre-pandemic period, SARS-CoV-2 was the most frequently detected virus, followed by influenza and RSV, similar to observations from a study in a German tertiary hospital, in which only SARS-CoV-2 screening was systematic^31^. By the 2024–2025 season, influenza had again become the predominant respiratory virus, consistent with surveillance findings from other countries^32^. Nevertheless, direct comparison of positivity rates between studies remains challenging because inclusion criteria, patient populations and testing strategies differ substantially.

Hospitalised patients were predominantly older men with multiple comorbidities, particularly cardiovascular disease, chronic lung disease and diabetes as described in previous studies^8,19,21,33^. Older patients generally had longer hospitalisations and higher in-hospital mortality rate, whereas ICU admission and respiratory support were more frequent in younger adults, probably reflecting differences in clinical management of frail elderly patients. Among laboratory-confirmed detections, influenza was generally associated with shorter hospitalisations, while RSV and hMPV more frequently required respiratory support. hMPV involved longer hospitalisations in younger adults. All these results are in agreement with recent studies in hospitalised adults^29,30,34^. The relatively longer hospital stays observed in some patients with rhinovirus detection who did not require ICU admission may reflect the selective testing of patients with greater clinical complexity or underlying chronic disease rather than purely greater intrinsic pathogenicity of rhinovirus, consistent with prospective studies showing that rhinovirus frequently affects older adults with multiple comorbidities^8,35^.

A major strength of this study was the integration of routinely collected administrative variables, severity outcomes and laboratory data for more than 100,000 ARI-related hospital visits over almost a decade. Unlike administrative databases alone, the linkage of patients’ characteristics and microbiological data improved pathogen ascertainment, while the availability of age- and time-specific catchment populations enabled calculation of population-based incidence rates. Such real-world datasets are becoming increasingly valuable for respiratory surveillance, disease burden estimation and evaluation of preventive interventions, particularly when combined with robust population denominators^12^. These data could also be used to identify early predictors of patient deterioration, in order to improve their care and the management of hospital beds during epidemics.

This study has several limitations. The data were collected as part of routine patient care rather than according to a research protocol and therefore depend on the completeness and accuracy of electronic health records. Comorbidities were identified from ICD-10 codes assigned during the ARI hospitalisation and may have been incompletely captured, while coding practices may also have evolved over time, particularly during the COVID-19 pandemic. Virological testing strategies changed considerably throughout the study period. Testing was mostly clinician-directed, with periods of systematic testing of SARS-CoV-2 even for asymptomatic patients. The possible use of triplex or panel tests to detect several viruses at once also increased the number of negative tests, as, for example, SARS-CoV-2 and RSV may also have been tested for a suspicion of influenza. This may result in under-ascertainment of some respiratory viruses and overestimation of others, limiting direct comparison of positivity rates across seasons and comparison with systematically tested cohorts. Consequently, the observed temporal trends reflect both virus circulation and changes in testing practices. Viral detection did not necessarily reflect active infection, as prolonged viral shedding can be detected, including in asymptomatic patients. In addition, comparisons between viruses were descriptive and were not adjusted for differences in patient characteristics or clinical management such as bed occupancies. The differences should therefore not be interpreted as causal. Finally, although HCL is the second largest university hospital in France, these findings originate from a single healthcare network and may not be fully generalisable to other healthcare settings.

In conclusion, this study provides a comprehensive description of nearly a decade of real-world data on adults with ARI in a large tertiary healthcare hospital. Beyond documenting changes in respiratory virus epidemiology before, during and after the COVID-19 pandemic, it illustrates how evolving diagnostic practices influence the interpretation of routinely collected surveillance data. By integrating administrative data, severity outcomes and laboratory information with population-based denominators, this cohort provides a platform for respiratory surveillance and further analyses of disease burden, severity and healthcare use. In particular, future analyses of these data could help identify predictive risk factors of severity, detect at-risk patients early and tailor adequate care.

## ETHICS

The research on data from the HCL received authorisation from the Commission Nationale de l’Informatique et des Libertés (CNIL) number 923273 on 2023/11/06 in accordance with the Declaration of Helsinki. Requirement for individual patient consent was waived in favour of an opt-out system whereby the details of the inclusion criteria for patients were published at least one month before their use and patients could therefore request to have their data excluded.

## DATA AVAILABILITY STATEMENT

The use of these data by was approved by CNIL. Due to legal and ethical restrictions related to the protection of personal health information, individual-level data are not publicly available.

## ACKNOWLEDGEMENTS

We would like to thank all the HCL staff who contributed to the preparation of the dataset, and all the patients who participated in the study.

## REFERENCES

1. Chen C, You Y, Du Y, et al. Global epidemiological trends in the incidence and deaths of acute respiratory infections from 1990 to 2021. Heliyon. 2024;10(16):e35841. doi:10.1016/j.heliyon.2024.e35841

2. Centre européen de prévention et de contrôle des maladies (Organisme et agence de l’UE), Alexakis, Leonidas, Barbezange, Cyril, et al. Integrated Respiratory Virus Surveillance Guidance for the EU/EEA. Office des publications de l’Union européenne; 2026. Accessed August 27, 2026. https://data.europa.eu/doi/10.2900/9293378

3. Nunes MC, Chauvel C, Raboni SM, et al. The Global Influenza Hospital Surveillance Network: A Multicountry Public Health Collaboration. Influenza Other Respir Viruses. 2025;19(3):e70091. doi:10.1111/irv.70091

4. Chow EJ, Uyeki TM, Chu HY. The effects of the COVID-19 pandemic on community respiratory virus activity. Nat Rev Microbiol. 2023;21(3):195–210. doi:10.1038/s41579-022-00807-9

5. Loubet P, Guitton S, Rolland S, et al. Characteristics of Human Metapneumovirus Infection Compared to Respiratory Syncytial Virus and Influenza Infections in Adults Hospitalized for Influenza-Like Illness in France, 2012-2022. J Infect Dis. 2025;232(Supplement_1):S93–S100. doi:10.1093/infdis/jiaf082

6. Muscatello DJ, Rose N, Paul KK, et al. Epidemiological comparison of emergency department presentations with seasonal influenza or COVID-19 and an outcome of intensive care admission or death: A population-based records linkage study in New South Wales, Australia. J Infect. 2024;89(6):106307. doi:10.1016/j.jinf.2024.106307

7. Piroth L, Cottenet J, Mariet AS, et al. Comparison of the characteristics, morbidity, and mortality of COVID-19 and seasonal influenza: a nationwide, population-based retrospective cohort study. Lancet Respir Med. 2021;9(3):251–259. doi:10.1016/S2213-2600(20)30527-0

8. Chaves SS, Castells VB, Mira-Iglesias A, et al. Rhinovirus/enterovirus contribution to respiratory-associated hospitalizations in adults during respiratory seasons in Spain: A 6-year prospective study. PloS One. 2026;21(4):e0347659. doi:10.1371/journal.pone.0347659

9. Lomholt FK, Møller KL, Nielsen J, Valentiner-Branth P, Vestergaard LS. Surveillance of severe acute respiratory infections using ICD-10 diagnosis codes and national electronic health records, Denmark, 2022 to 2024. Eurosurveillance. 2025;30(33):2400801. doi:10.2807/1560-7917.ES.2025.30.33.2400801

10. INSEE. Dossier complet - Intercommunalité - Métropole de Lyon. Published online August 9, 2022. https://www.insee.fr/fr/statistiques/2011101?geo=EPCI-200046977#chiffre-cle-1

11. Stellrecht KA, Cimino JL, Wilson LI, Maceira VP, Butt SA. Panther Fusion® Respiratory Virus Assays for the detection of influenza and other respiratory viruses. J Clin Virol. 2019;121:104204. doi:10.1016/j.jcv.2019.104204

12. Shirreff G, Chauvel C, Casalegno JS, et al. Estimation of hospital catchment populations using data on patient hospital use in France. medRxiv. Preprint posted online April 29, 2026:2026.04.28.26351911. doi:10.64898/2026.04.28.26351911

13. Ulm K. SIMPLE METHOD TO CALCULATE THE CONFIDENCE INTERVAL OF A STANDARDIZED MORTALITY RATIO (SMR). Am J Epidemiol. 1990;131(2):373–375. doi:10.1093/oxfordjournals.aje.a115507

14. Williams B, Mancia G, Spiering W, et al. 2018 ESC/ESH Guidelines for the management of arterial hypertension: The Task Force for the management of arterial hypertension of the European Society of Cardiology (ESC) and the European Society of Hypertension (ESH). Eur Heart J. 2018;39(33):3021–3104. doi:10.1093/eurheartj/ehy339

15. Mills KT, Bundy JD, Kelly TN, et al. Global Disparities of Hypertension Prevalence and Control: A Systematic Analysis of Population-based Studies from 90 Countries. Circulation. 2016;134(6):441–450. doi:10.1161/CIRCULATIONAHA.115.018912

16. Chow CK, Teo KK, Rangarajan S, et al. Prevalence, Awareness, Treatment, and Control of Hypertension in Rural and Urban Communities in High-, Middle-, and Low-Income Countries. JAMA. 2013;310(9):959–968. doi:10.1001/jama.2013.184182

17. Benjamini Y, Hochberg Y. Controlling the False Discovery Rate: A Practical and Powerful Approach to Multiple Testing. J R Stat Soc Ser B Methodol. 1995;57(1):289–300.

18. R Core Team. R: A Language and Environment for Statistical Computing. R Foundation for Statistical Computing. Published online 2022. https://www.R-project.org

19. Loubet P, Lenzi N, Valette M, et al. Clinical characteristics and outcome of respiratory syncytial virus infection among adults hospitalized with influenza-like illness in France. Clin Microbiol Infect Off Publ Eur Soc Clin Microbiol Infect Dis. 2017;23(4):253–259. doi:10.1016/j.cmi.2016.11.014

20. Loubet P, Mathieu P, Lenzi N, et al. Characteristics of human metapneumovirus infection in adults hospitalized for community-acquired influenza-like illness in France, 2012–2018: a retrospective observational study. Clin Microbiol Infect. 2021;27(1):127.e1-127.e6. doi:10.1016/j.cmi.2020.04.005

21. Bénézit F, Loubet P, Galtier F, et al. Non-influenza respiratory viruses in adult patients admitted with influenza-like illness: a 3-year prospective multicenter study. Infection. 2020;48(4):489–495. doi:10.1007/s15010-019-01388-1

22. Fonseca MJ, Bratković S, Pedersen M, et al. Incidence of respiratory syncytial virus and influenza: A Danish nationwide cohort study. Hum Vaccines Immunother. 2026;22(1):2638638. doi:10.1080/21645515.2026.2638638

23. Stojanovic Z, Gonçalves-Carvalho F, Marín A, et al. Advances in diagnostic tools for respiratory tract infections: from tuberculosis to COVID-19 – changing paradigms? ERJ Open Res. 2022;8(3):00113–02022. doi:10.1183/23120541.00113-2022

24. Egeskov-Cavling AM, Johannesen CK, Lindegaard B, Fischer TK, PROMISE Investigators. Underreporting and Misclassification of Respiratory Syncytial Virus–Coded Hospitalization Among Adults in Denmark Between 2015–2016 and 2017–2018. J Infect Dis. 2024;229(Supplement_1):S78–S83. doi:10.1093/infdis/jiad415

25. Li Y, Kulkarni D, Begier E, et al. Adjusting for Case Under-Ascertainment in Estimating RSV Hospitalisation Burden of Older Adults in High-Income Countries: a Systematic Review and Modelling Study. Infect Dis Ther. 2023;12(4):1137–1149. doi:10.1007/s40121-023-00792-3

26. Moriyama M, Hugentobler WJ, Iwasaki A. Seasonality of Respiratory Viral Infections. Annu Rev Virol. 2020;7(Volume 7, 2020):83–101. doi:10.1146/annurev-virology-012420-022445

27. Decker SR da R, Wolf JM, Pille A, et al. Temporal trends in respiratory pathogens following the COVID-19 pandemic and climate variables: A unicentric retrospective evaluation of 24 pathogens in a temperate subtropical region. J Med Virol. 2024;96(7):e29797. doi:10.1002/jmv.29797

28. Domnich A, Panatto D, Paolozzi V, et al. Epidemiology and Burden of Human Metapneumovirus Among Italian Adults in Outpatient and Inpatient Settings, 2014–2025. Influenza Other Respir Viruses. 2025;19(10):e70175. doi:10.1111/irv.70175

29. Sanz-Muñoz I, Galindo-Carretero S, Toquero-Asensio M, et al. Characteristics and Risk Factors Associated With Hospitalization and Severe Disease in Patients Aged ≥ 60 Years With Human Metapneumovirus Infection Over Three Consecutive Seasons (2021–2024) in Valladolid, Spain. The VALLAVIRUS Study. Influenza Other Respir Viruses. 2026;20(7):e70294. doi:10.1111/irv.70294

30. Sanz-Muñoz I, Iglesia-Aparicio R, López-Gonzalo C, et al. Incidence, Complications, and Quality of Life in Adults ≥ 60 Years Old With Laboratory-Confirmed RSV Infections Attended in Primary Care and in Hospitals: A Study Over Three Seasons in Valladolid, Spain. Influenza Other Respir Viruses. 2026;20(7):e70295. doi:10.1111/irv.70295

31. Mees J, Eisenmann M, Höhn A, et al. Frequency, lethality, and demographic trends of acute respiratory viruses in hospitalized patients: insights from a German tertiary care hospital from july 2022 to april 2023. BMC Infect Dis. 2025;25(1):1110. doi:10.1186/s12879-025-11521-1

32. Aguilar Figueroa D, Pérez-Gimeno G, Núñez O, Monge S. Burden of Severe Disease Associated With Influenza, SARS-CoV-2 and RSV in Spain During the 2024–2025 Winter Season. Influenza Other Respir Viruses. 2025;19(11):e70190. doi:10.1111/irv.70190

33. Puggina A, Dovizio M, Domnich A, et al. Demographics and clinical burden of disease among RSV-hospitalized older adults in Italy: A retrospective cohort study. Hum Vaccines Immunother. 2025;21(1):2479334. doi:10.1080/21645515.2025.2479334

34. Correcher-Martínez E, Muñoz-Quiles C, López-Lacort M, et al. Severity and Cost of Respiratory Syncytial Virus Versus Influenza in Hospitalized Adults in Spain. Influenza Other Respir Viruses. 2026;20(4):e70254. doi:10.1111/irv.70254

35. Revilla AM, Raboni SM, Giamberardino HIG, et al. Clinical Characteristics and Severity of Rhinovirus/Enterovirus–Associated Hospitalizations: A Multicountry Analysis From the Global Influenza Hospital Surveillance Network, 2017–2024. Open Forum Infect Dis. 2026;13(4):ofag146. doi:10.1093/ofid/ofag146

