## Supplementary data for "Epidemiology of acute respiratory infections in adult patients in Hospices Civils de Lyon, France, between October 2016 and April 2025"

*Supplementary Table 1 : ICD-10 codes used in the study for patient inclusion by diagnostic codes and identification of comorbidities*

|  | **ICD-10 codes** |
| --- | --- |
| **Acute respiratory infection related admission (patient inclusion)** | J09-J18, J20-J22, J440, J85, J86, A481, B012, B052, B250, B371, B380, B390, B400, B410, B420, B440, B450, B460, B583, B59, P23, U0710, U0711 |
| **Comorbidities** |  |
| Cardiovascular disease | I05–I09, I10, I25, I310, I311, I420, I481, I482, I489, I501  I5020, I5022, I5023, I5030, I5032, I5033, I5040, I5042, I5043, I50810, I50812, I50813, I50814, I5082, I5083,  I5084, I5089, I509, I65, I66, I67, I69, I70, I73, Q20–Q28, |
| Hypertension | I10 |
| Chronic lung disease | J41, J42, J43, J44, J961, E840 |
| Diabetes | E08, E09, E10, E11, E13, E14, |
| Neoplasm | C00–C96, Z511, Z512 |
| Neurological disease | G10–G12, G20–G21, G30–G31, G35, G40, G43, G70–G71, G80–G83 |
| Renal disease | N03, N11, N18, N18, Z49, Z992 |
| Immunological disease | B20, B21, B22, B23, B24, D648, D70, D71, D728, D758, D762, D80, D81, D82, D83, D84, D86, D89, Z94, Z7952 |
| Obesity | E660, E661, E662, E668, E669 |
| Hemoglobinopathies | D55–D61, D65, D66, D67, D680, D681, D682, D750, D751 |
| Liver disease | B18, B199, K70, K713-K717, K721, K729, K73, K74, K760, K766, K811 |
| Rheumatologic disease | M05, M06, M08, M30-M35, M45 |
| Asthma | J45 |
| Tuberculosis | A150–A199 |

Supplementary Table 2: Hospital visits and inclusions by ICD-10 code or viral testing over the study seasons

|  | **Emergency only** | | | **Hospitalisation** | | |
| --- | --- | --- | --- | --- | --- | --- |
|  | **Overall** | **ICD-10***^1^* | **Viral testing***^1^* | **Overall***^1^* | **ICD-10***^1^* | **Viral testing***^1^* |
| **Overall**  N = 103,011 | 19,948 | 11,765 (59%) | 8,183 (41%) | 83,063 | 52,928 (64%) | 30,135 (36%) |
| **Season** |  |  |  |  |  |  |
| 2016-2017 | 1,262 | 1,193 (95%) | 69 (5.5%) | 7,083 | 6,158 (87%) | 925 (13%) |
| 2017-2018 | 1,590 | 1,510 (95%) | 80 (5.0%) | 7,788 | 6,710 (86%) | 1,078 (14%) |
| 2018-2019 | 1,268 | 1,158 (91%) | 110 (8.7%) | 7,789 | 6,639 (85%) | 1,150 (15%) |
| 2019-2020* | 1,885 | 1,483 (79%) | 402 (21%) | 8,367 | 6,258 (75%) | 2,109 (25%) |
| 2020-2021* | 2,584 | 1,436 (56%) | 1,148 (44%) | 12,214 | 6,207 (51%) | 6,007 (49%) |
| 2021-2022* | 3,596 | 1,380 (38%) | 2,216 (62%) | 11,948 | 5,044 (42%) | 6,904 (58%) |
| 2022-2023 | 2,938 | 1,239 (42%) | 1,699 (58%) | 10,542 | 5,301 (50%) | 5,241 (50%) |
| 2023-2024 | 2,724 | 1,339 (49%) | 1,385 (51%) | 10,112 | 6,065 (60%) | 4,047 (40%) |
| 2024-2025 | 2,101 | 1,027 (49%) | 1,074 (51%) | 7,220 | 4,546 (63%) | 2,674 (37%) |

*^1^ n (%); *seasons during the COVID-19 pandemic*

*Supplementary Figure 1: Number of ARI-related hospital visits with at least one viral test for influenza A, influenza B, SARS-CoV-2, RSV, hMPV, rhinovirus and other viruses (PIV, ADV, hCoV, enterovirus and enterovirus/rhinovirus) by epidemic season*

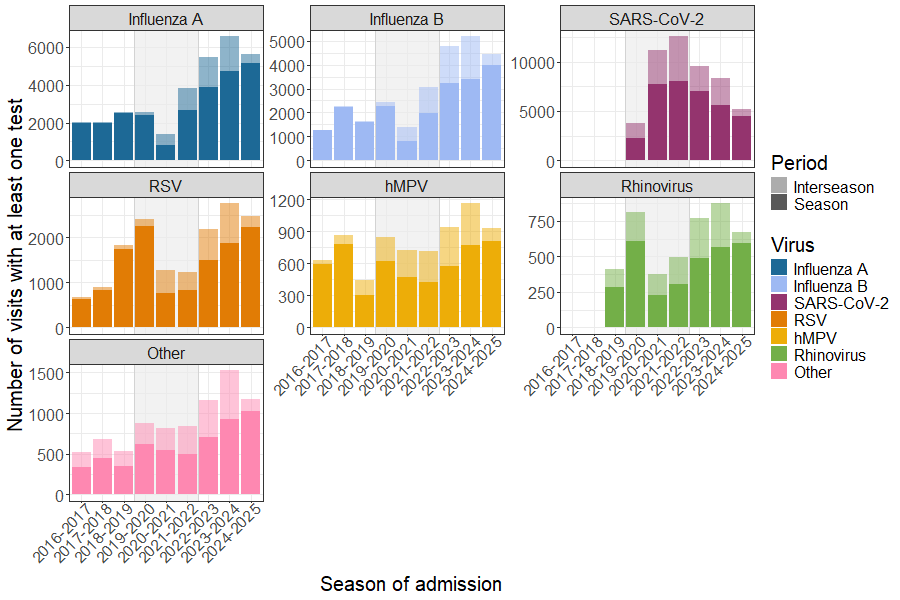
*Note: rhinovirus not tested before November 2018; Grey shading: COVID-19 pandemic seasons*

Supplementary Figure 2: Positivity rates of influenza A, influenza B, SARS-CoV-2, RSV, hMPV and rhinovirus tests by season and age group in patients with acute respiratory infections

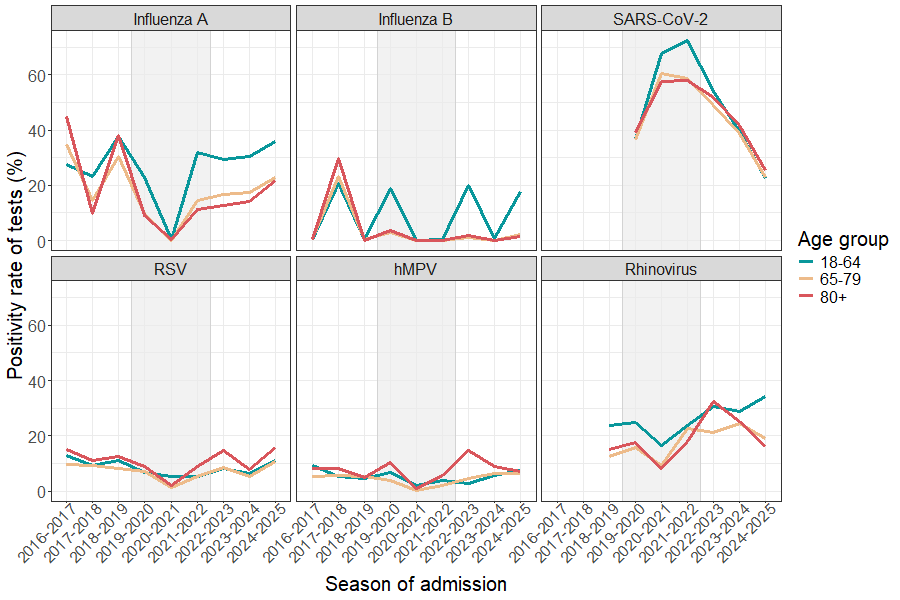
*Grey shading: COVID-19 pandemic seasons*

*Supplementary Figure 3: Number of visits with at least one positive viral test for adenovirus (ADV), enterovirus/rhinovirus (EV/RV), human coronavirus (hCoV) and parainfluenza virus (PIV) by epidemic season*

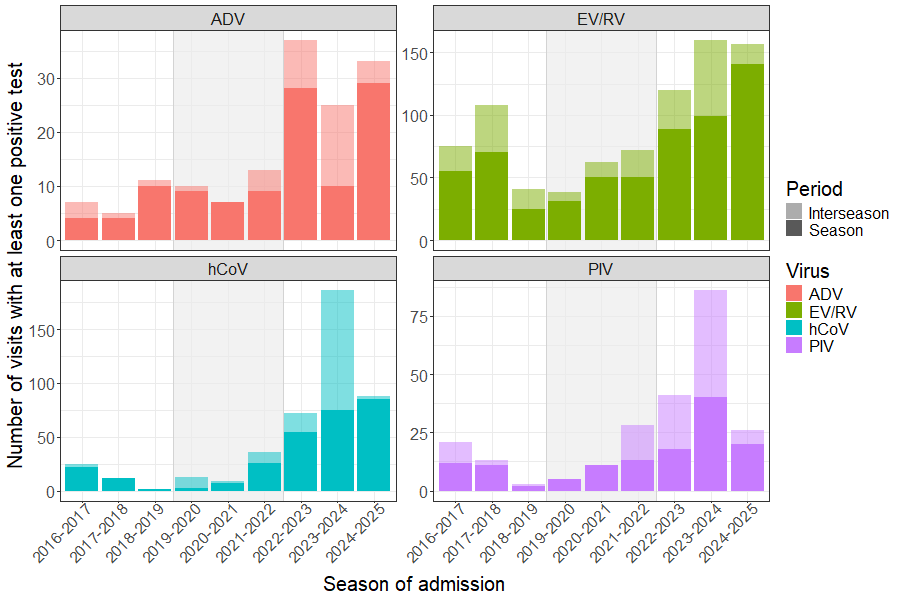

*Grey shading: COVID-19 pandemic seasons*

Supplementary Table 3: Comorbidities of hospitalised patients with ARI overall, by period related to the COVID-19 pandemic and by age group

| **Comorbidity, n (%)** | **Overall** | **COVID-19 pandemic period** | | | | | | | | **Age group** | | | | | |
| --- | --- | --- | --- | --- | --- | --- | --- | --- | --- | --- | --- | --- | --- | --- | --- |
|  |  | **Before** | | **During** | | **After** | |  | | **18-64** | **65-79** | | **≥80** | |  |
|  | N = 83,063 | N = 22,660 | | N = 32,529 | | N = 27,874 | | **p-value** | | N = 28,781 | N = 25,331 | | N = 28,951 | | **p-value** |
| **Cardiovascular disease** | 39,176 (47) | 10,547 (47) | | 14,865 (46) | | 13,764 (49) | | <0.001 | | 6,568 (23) | 13,641 (54) | | 18,967 (66) | | <0.001 |
| **Cardiovascular disease excluding hypertension** | 25,335 (31) | 7,746 (34) | | 8,968 (28) | | 8,621 (31) | | <0.001 | | 3,742 (13) | 8,515 (34) | | 13,078 (45) | | <0.001 |
| **Chronic lung disease** | 17,412 (21) | 6,292 (28) | | 5,484 (17) | | 5,636 (20) | | <0.001 | | 5,090 (18) | 6,974 (28) | | 5,348 (18) | | <0.001 |
| **Diabetes** | 16,284 (20) | 4,234 (19) | | 6,533 (20) | | 5,517 (20) | | <0.001 | | 3,640 (13) | 6,701 (26) | | 5,943 (21) | | <0.001 |
| **Neoplasm** | 16,011 (19) | 4,669 (21) | | 5,724 (18) | | 5,618 (20) | | <0.001 | | 5,281 (18) | 6,936 (27) | | 3,794 (13) | | <0.001 |
| **Neurological disease** | 14,201 (17) | 4,003 (18) | | 5,140 (16) | | 5,058 (18) | | <0.001 | | 3,133 (11) | 3,569 (14) | | 7,499 (26) | | <0.001 |
| **Renal disease** | 9,756 (12) | 2,593 (11) | | 3,382 (10) | | 3,781 (14) | | <0.001 | | 1,688 (5.9) | 3,264 (13) | | 4,804 (17) | | <0.001 |
| **Immunological disease** | 7,872 (9.5) | 1,705 (7.5) | | 3,364 (10) | | 2,803 (10) | | <0.001 | | 3,808 (13) | 2,504 (9.9) | | 1,560 (5.4) | | <0.001 |
| **Obesity** | 7,074 (8.5) | 1,518 (6.7) | | 3,572 (11) | | 1,984 (7.1) | | <0.001 | | 2,982 (10) | 2,652 (10) | | 1,440 (5.0) | | <0.001 |
| **Hemoglobinopathies** | 3,393 (4.1) | 952 (4.2) | | 1,153 (3.5) | | 1,288 (4.6) | | <0.001 | | 1,969 (6.8) | 1,061 (4.2) | | 363 (1.3) | | <0.001 |
| **Cirrhosis, liver disease** | 2,937 (3.5) | 780 (3.4) | | 1,115 (3.4) | | 1,042 (3.7) | | 0.08 | | 1,465 (5.1) | 1,093 (4.3) | | 379 (1.3) | | <0.001 |
| **Rheumatologic disease** | 2,460 (3.0) | 700 (3.1) | | 911 (2.8) | | 849 (3.0) | | 0.086 | | 832 (2.9) | 820 (3.2) | | 808 (2.8) | | 0.006 |
| **Asthma** | 2,267 (2.7) | 573 (2.5) | | 886 (2.7) | | 808 (2.9) | | 0.04 | | 1,007 (3.5) | 650 (2.6) | | 610 (2.1) | | <0.001 |
| **Tuberculosis** | 211 (0.3) | 72 (0.3) | | 71 (0.2) | | 68 (0.2) | | 0.068 | | 129 (0.4) | 52 (0.2) | | 30 (0.1) | | <0.001 |

*Supplementary Table 4: Characteristics of hospitalised patients with acute respiratory infections and their severity outcomes overall and by age group (18-64, 65-79 or ≥80)*

| **Characteristics** | **Overall** N = 83,063 | **Age group** | | | |
| --- | --- | --- | --- | --- | --- |
|  |  | **18-64** N = 28,781 | **65-79** N = 25,331 | **≥80** N = 28,951 | **p-value** |
| **Length of first stay in days, median (IQR)** | 9 (13) | 8 (12) | 10 (14) | 10 (12) | <0.001 |
| **Readmission within 30 days, n (%)** | 5,928 (7.1) | 2,004 (7.0) | 2,034 (8.0) | 1,890 (6.5) | <0.001 |
| **Women, n (%)** | 37,285 (45) | 12,318 (43) | 9,633 (38) | 15,334 (53) | <0.001 |
| **Emergency department visit, n (%)** | 49,819 (60) | 13,501 (47) | 14,111 (56) | 22,207 (77) | <0.001 |
| **Type of discharge, n (%)** |  |  |  |  | <0.001 |
| In-hospital death | 9,708 (12) | 1,973 (6.9) | 3,242 (13) | 4,493 (16) | <0.001 |
| Home | 51,117 (62) | 20,940 (73) | 15,248 (60) | 14,929 (52) | <0.001 |
| Transfer | 22,238 (27) | 5,868 (20) | 6,841 (27) | 9,529 (33) | <0.001 |
| **Laboratory-confirmed**  **viral infection, n (%)** | 30,135 (36) | 10,955 (38) | 8,869 (35) | 10,311 (36) | <0.001 |
| **Laboratory-confirmed**  **viral co-infection, n (%)** | 781 (0.9) | 361 (1.3) | 245 (1.0) | 175 (0.6) | <0.001 |
| **Number of comorbidities, n (%)** |  |  |  |  | <0.001 |
| None | 16,287 (20) | 8,574 (30) | 3,265 (13) | 4,448 (15) | <0.001 |
| One | 19,424 (23) | 8,741 (30) | 5,563 (22) | 5,120 (18) | <0.001 |
| One, excluding hypertension | 27,284 (33) | 10,109 (35) | 7,924 (31) | 9,251 (32) | <0.001 |
| At least two | 47,352 (57) | 11,466 (40) | 16,503 (65) | 19,383 (67) | <0.001 |
| At least two, excluding hypertension | 36,731 (44) | 9,613 (33) | 13,367 (53) | 13,751 (47) | <0.001 |
| **ICU, n (%)** | 21,094 (25) | 9,617 (33) | 8,124 (32) | 3,353 (12) | <0.001 |
| **Oxygen supplementation, n (%)** | 16,617 (20) | 7,458 (26) | 6,422 (25) | 2,737 (9.5) | <0.001 |
| **Non-invasive mechanical ventilation, n (%)** | 12,157 (15) | 5,164 (18) | 4,950 (20) | 2,043 (7.1) | <0.001 |
| **Invasive mechanical ventilation, n (%)** | 10,283 (12) | 5,269 (18) | 4,077 (16) | 937 (3.2) | <0.001 |
| **ECMO, n (%)** | 310 (0.4) | 261 (0.9) | 49 (0.2) | 0 (0) | <0.001 |
| *ICU: intensive care unit; ECMO: extracorporeal membrane oxygenation* | | | | | |
